# Molecular Genetic Analysis of SARS-CoV-2 Lineages in Armenia

**DOI:** 10.1101/2021.06.19.21259172

**Authors:** Diana Avetyan, Siras Hakobyan, Maria Nikoghosyan, Gisane Khachatryan, Tamara Sirunyan, Nelli Muradyan, Roksana Zakharyan, Andranik Chavushyan, Hovsep Ghazaryan, Ani Melkonyan, Ani Stepanyan, Varduhi Hayrapetyan, Sofi Atshemyan, Gevorg Martirosyan, Gayane Melik-Andreasyan, Shushan Sargsyan, Armine Ghazazyan, Naira Aleksanyan, Lilit Nersisyan, Arsen Arakelyan

## Abstract

Sequencing of SARS-CoV-2 provides essential information into viral evolution, transmission, and epidemiology. Short-read next-generation sequencing platforms are currently the gold-standard approaches characterized by the highest accuracy. Meanwhile, Oxford Nanopore’s long-read sequencing devices show great promise, offering comparable accuracy, fast turnaround time, and reduced cost. In this study, we performed whole-genome sequencing and molecular-genetic characterization of SARS-CoV-2 from clinical specimens using an amplicon-based nanopore sequencing approach. Lineage and phylogenetic analysis identified the most prevalent lineages at different time points (B.1.1.163, B.1.1.208, B.1.1, and since March 2021 - B.1.1.7). In addition, we evaluated the possible effect of identified mutations on the efficacy of recommended primers and probes used for PCR detection of SARS-CoV-2. In summary, a high-quality SARS-CoV-2 genome can be acquired by nanopore sequencing and it can serve as an efficient and affordable alternative to short-read next-generation sequencing and be used for epidemiologic surveillance and molecular-genetic analyses of the virus.

## Introduction

Severe acute respiratory syndrome coronavirus-2 (SARS-CoV-2) was first identified in China, in the city of Wuhan, in December 2019, which causes novel coronavirus pneumonia COVID-19 [1]. The complete genome sequence was published in January 2020 [2–4], which led to the development of real-time reverse transcription-polymerase chain reaction (qRT-PCR) assays for SARS-CoV-2 detection that have served as the diagnostic standard during the ongoing COVID-19 pandemic [5].

Since then, whole-genome sequencing is used for the evolutionary analysis of viruses, monitoring of circulating genetic lineages, and identifying signs of adaptation to hosts, which have important implications for treatment and vaccine development [6–8]. The Illumina short-read whole-genome sequencing platforms enabled accurate sequence determination and are considered as a standard for pathogen genomics, and are currently the method of choice for SARS-CoV-2 sequencing [9]. However, whole-genome sequencing essential for public health in epidemiological monitoring and surveillance of viral pathogens is still challenging in many countries with poor technical resources. Thus, for rapid, easy, reproducible, and affordable SARS-CoV-2 genomic sequencing data an alternative protocol for nanopore sequencing has been developed and shown to have comparable accuracy with Illumina [10,11]. The advantage of nanopore sequencing is its relatively low cost and speed, minimal requirements of infrastructure, simple technical implementation, however, still certain concerns about sequencing accuracy are discussed [12,13]. A specific amplification-based enrichment was optimized for ONT, which can generate sufficient quantities of the genetic material needed for sequencing. Recent studies provide evidence on accurate consensus-level SARS-CoV-2 sequencing using ONT [14,15].

Here we applied a modified ARTIC protocol for ONT MinION to perform whole genome sequencing and molecular-genetic characterization of SARS-CoV-2 viruses from clinical specimens collected in Armenia.

## Material and methods

### Samples

Thirty-six RNA samples isolated from nasopharyngeal swabs were obtained from the National Center for Disease Control and Prevention, Ministry of Health RA (NCDC). These samples were selected from the batches of COVID-19 positive samples tested at NCDC on 22th and 29th January and 18 March 2021 (twelve samples on each day). SARS-Cov-2 PCR testing was performed using Real-Time PCR Detection Kit for COVID-19 Coronavirus CE-IVD kit (Biotech & Biomedicine (Shenyang) Group Ltd., China) targeting ORF1ab and N genes. Samples were selected based on the viral RNA load as measured by Ct values between 18-35 (Supplemental Table S1) for both targets.

Automated RNA isolation was performed with Maxwell RSC Instrument using Maxwell RSC Viral Total Nucleic Acid Purification Kit (Promega Corporation Inc, US).

### Nanopore sequencing

Nanopore sequencing was performed according to “nCoV-2019 sequencing protocol v3 (LoCost) V.3” [16] based on ARTIC SARS-CoV-2 sequencing protocol with ARTIC nCoV-2019 V3 PCR panel [17,18].

#### cDNA generation

RNA samples were directly used for the first-strand synthesis using the LunaScript RT SuperMix Kit (New England Biolabs, USA) with random hexamer and oligo-dT primers. Briefly, 8 μL RNA were mixed with 2 μL LunaScript RT SuperMix (5X) and were placed in a thermocycler and incubated 2 minutes at 25°C, followed by 10 minutes at 55°C and 1 minute at 95°C and cooling to 4°C. cDNAs were immediately used in subsequent steps.

#### Amplicon generation

Primer pairs from the ARTIC V3 primer scheme were used to amplify amplicons in cDNA [19]. Two Multiplex PCR reactions were performed with 2.5 μL cDNA, 12.5 μL Q5 Hot Start High-Fidelity 2× Master Mix (New England Biolabs, USA), and 4 μL ARTIC V3 pool 1 (10 μM) or 4 μL ARTIC V3 pool 2 (10 μM). PCR cycling conditions were: 98°C for 30 secs followed by 35 cycles of 98°C for 15 secs, 65°C for 5 mins and hold at 4°C. The amplified products were purified with an 0.4x volume of AMPure XP beads (Beckman Coulter, USA) to exclude small nonspecific fragments.

#### Barcoding and library preparation

The purified PCR amplicons were treated with NEBNext End repair/dA-tailing Module (New England Biolabs, USA) and were barcoded with native barcodes and sequencing adapters (EXP-NBD104 kit Oxford Nanopore Technologies, UK). 12 samples were multiplexed in each sequencing run.

#### Nanopore sequencing

After priming the flow cell, 15 ng of the final sequencing library diluted to a final volume of 75 μL was loaded. Following the ligation sequencing kit (SQK-LSK109, Oxford Nanopore Technologies, UK) protocol, MinION Mk1B was used to perform genome sequencing in an FLO-MINSP6 R 9.4.1 flow cell for 3-6 hours. The sequencing coverage is presented in Figure 1.

**Figure 1.**
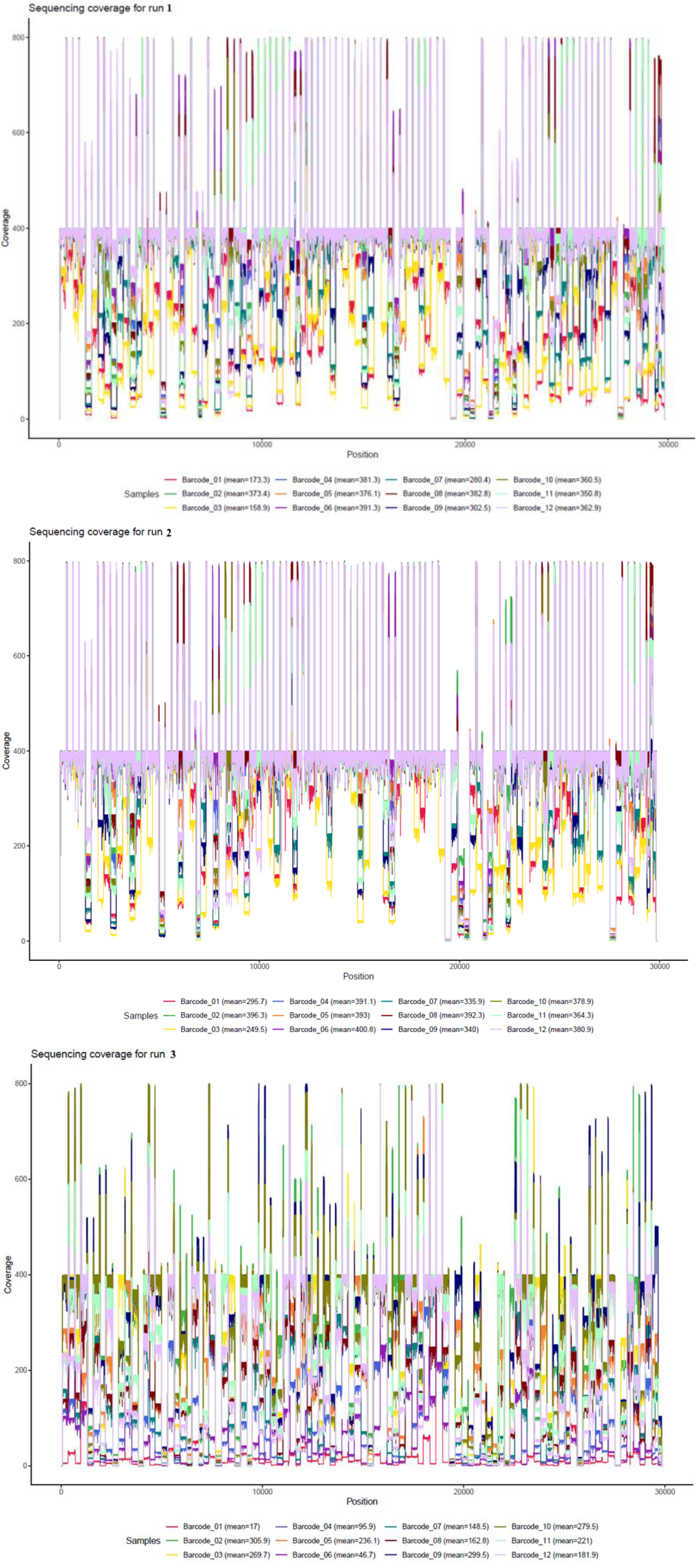
The sequencing coverage for each sample. The order of the positions corresponds to the reference genome (MN908947.3).

#### Data preprocessing, demultiplexing, and alignment

Read base calling and demultiplexing were performed using *Guppy* (4.0.14). The obtained FASTQ files were filtered and reads with length 400-700b were selected using ARTIC pipeline (release 1.1.0) [20]. The downstream analyses were performed using the nanopolish workflow implemented in the ARTIC pipeline [21]. The pipeline includes an alignment to the hCoV-19/Wuhan/WIV04 reference genome with minimap2 (2.17-r941) [22] followed by variant calling and consensus-building. The positions in consensus genomes with coverage lower than 20 were masked with “N” bases. For subsequent analyses “N” in genomes were substituted with corresponding nucleotides of the reference genome as described in Li et al (2021) [18].

### Phylogeny and variant analysis

Variant analysis and functional annotation were performed using the CoVsurver mutation app [23], CoV-GLUE [24], and CorGAT [25] tools.

Multiple alignment was performed with the MAFFT version 7 tool. A phylogenetic tree was built using “dist.alignment”, “phylogram”, “dendextend” and “ggtree” R packages. Clade and lineage analysis was performed using the pangolin tool [26].

To construct the phylogenetic tree, we have used our consensus sequences with low-quality bases substituted with reference. We also included three additional samples from Armenia submitted to GISAID [27,28] on 12 September 2020 (accessions: EPI_ISL_683451, EPI_ISL_683450, EPI_ISL_68344).

### Data availability

Consensus FASTA files were deposited in the GISAID EpiCoV database (accessions:

EPI_ISL_1139153, EPI_ISL_1139152, EPI_ISL_1139151, EPI_ISL_1139150,

EPI_ISL_1139157, EPI_ISL_1139156, EPI_ISL_1139155, EPI_ISL_1139154,

EPI_ISL_1718295, EPI_ISL_1718294, EPI_ISL_1718293, EPI_ISL_1718292,

EPI_ISL_1718299, EPI_ISL_1718298, EPI_ISL_1718297, EPI_ISL_1718296,

EPI_ISL_1139149, EPI_ISL_1139148, EPI_ISL_1139147, EPI_ISL_1718303,

EPI_ISL_1718302, EPI_ISL_1718301, EPI_ISL_1718300, EPI_ISL_1139146,

EPI_ISL_1718305, EPI_ISL_1718304, EPI_ISL_1718291, EPI_ISL_1718290,

EPI_ISL_1718289, EPI_ISL_1718284, EPI_ISL_1718283, EPI_ISL_1718282,

EPI_ISL_1718288, EPI_ISL_1718287, EPI_ISL_1718286, EPI_ISL_1718285).

## Results

This paper presents the results of nanopore sequencing of 36 SARS-CoV-2 isolated from clinical samples in Armenia in January and March 2021. The average sequencing coverage was 320x. Before proceeding with analyses we performed sequence quality assessment using the Nextclade web app. We noticed considerable variations in consensus qualities, especially in terms of the number of called ambiguous bases. For further analyses, we replaced “N” bases in consensus FASTA sequences with corresponding nucleotides from the reference genome as described in Li et al (2021) [10].

Next, we compared the obtained sequences with the Wuhan genome reference sequence (hCoV-19/Wuhan/WIV04). In samples obtained during January, overall 63 mutations were detected with varying frequencies. All samples contained the D614G mutation, which became a dominant mutation from March 2020 and replaced the original virus strain. Along with D614G, eighteen other mutations were also detected at high frequency (75-100%). Based on the mutation signature CoVsurver assigned all studied samples to the clade GR (C241T, C3037T, A23403G, G28882A includes S-D614G + N-G204R). On average 11±2 mutations were identified in each sample with 2±1 mutations reported as unique by the CoVsurver mutation app (Supplemental table S2). The unique mutations were mostly located in the non-structural protein genes (NS3_S220C, NS6_E55Q, NSP13_I35V, NSP2_L270F, NSP3_A1769S, NSP6_I189L), while two mutations were found in the N gene (N_Q160R, N_R203E), and one in the S gene (Spike_N460I). Fourteen mutations (1 unique) were identified as under selection (13 positive and 1 negative). T cell-mediated immunity target epitope prediction [25,29] showed that 74 mutations (7 unique) were located in epitopes that likely have the affinity of HLA class I and II molecules. Two mutations were identified with high frequency in SARS-CoV-2 epitopes restricted by HLA-A*24:02, one of the most prominent HLA class I alleles.

PANGO lineage analysis assigned twenty-one samples to B.1.1.163 (Russian) lineage; 2 samples were assigned B.1.1.208 and 1 sample was assigned to B.1.1 lineage. Interestingly, from three samples from Armenia submitted to GISAID in September 2020 two belonged to B.4 (Iranian) lineage, which was almost entirely replaced by B.1 lineages globally (Figure 2). The B.1.1.163 lineage was the most frequent in Russia (Russia 36.0%, United Kingdom 20.0%, United States of America 7.0%, Armenia 7.0%, Japan 6.0%), which was the first country Armenia opened borders with. No lineages representing variants of concern (B.1.1.7, B.1.351, and P.1) were detected among the January samples, however, we observed several mutations characteristic to B.1.1.7: Spike P681H in 6 samples, as well as Spike S982A and N S235F, each in one sample.

**Figure 2.**
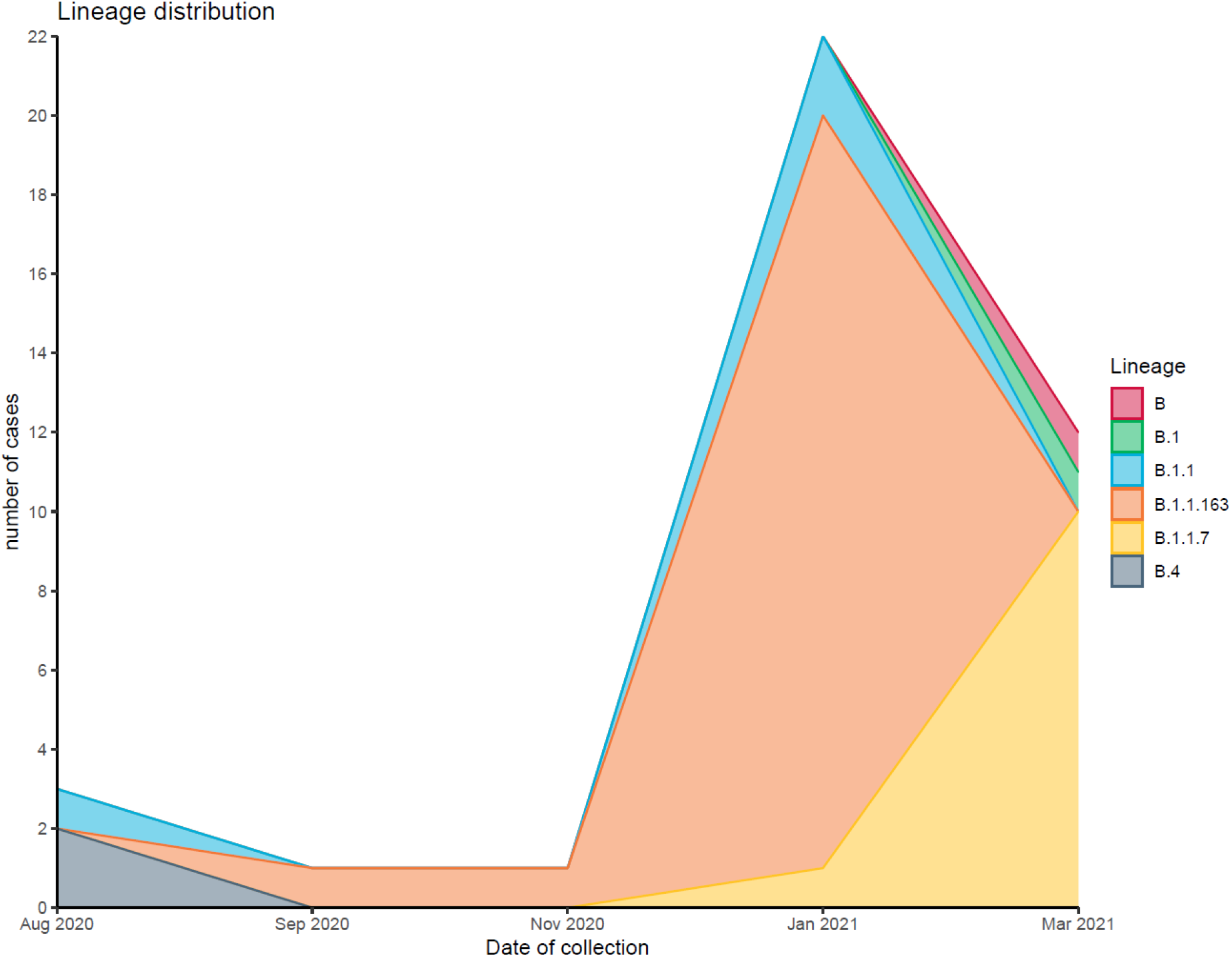
SARS-CoV-2 virus lineage frequencies. This graph shows the changing of the frequencies of SARS-CoV-2 lineages in Armenia. Samples were collected from August 2020 to March 2021.

The lineage composition has been changed dramatically in clinical samples collected in March. PANGO analysis assigned all sequences to B.1.1.7 Alfa (UK) lineage with characteristic N501Y, A570D, P681H, and NS8 Q27stop mutations found in all samples. No E484K/Q “immune escape” mutation was identified.

Phylogenetic analysis of 264 B.1.1.163 lineage sequences has placed 24 samples collected in January in the cluster along with sequences from Europe and Russia suggesting the possible transmission route for B.1.1.163 to Armenia. In contrast, analysis of B.1.1.7 sequences from 12 Armenian samples along with 448 sequences clustered them on different phylogenetic tree branches, which indicates multiple routes of transmission of this lineage to Armenia (Figure 3).

**Figure 3.**
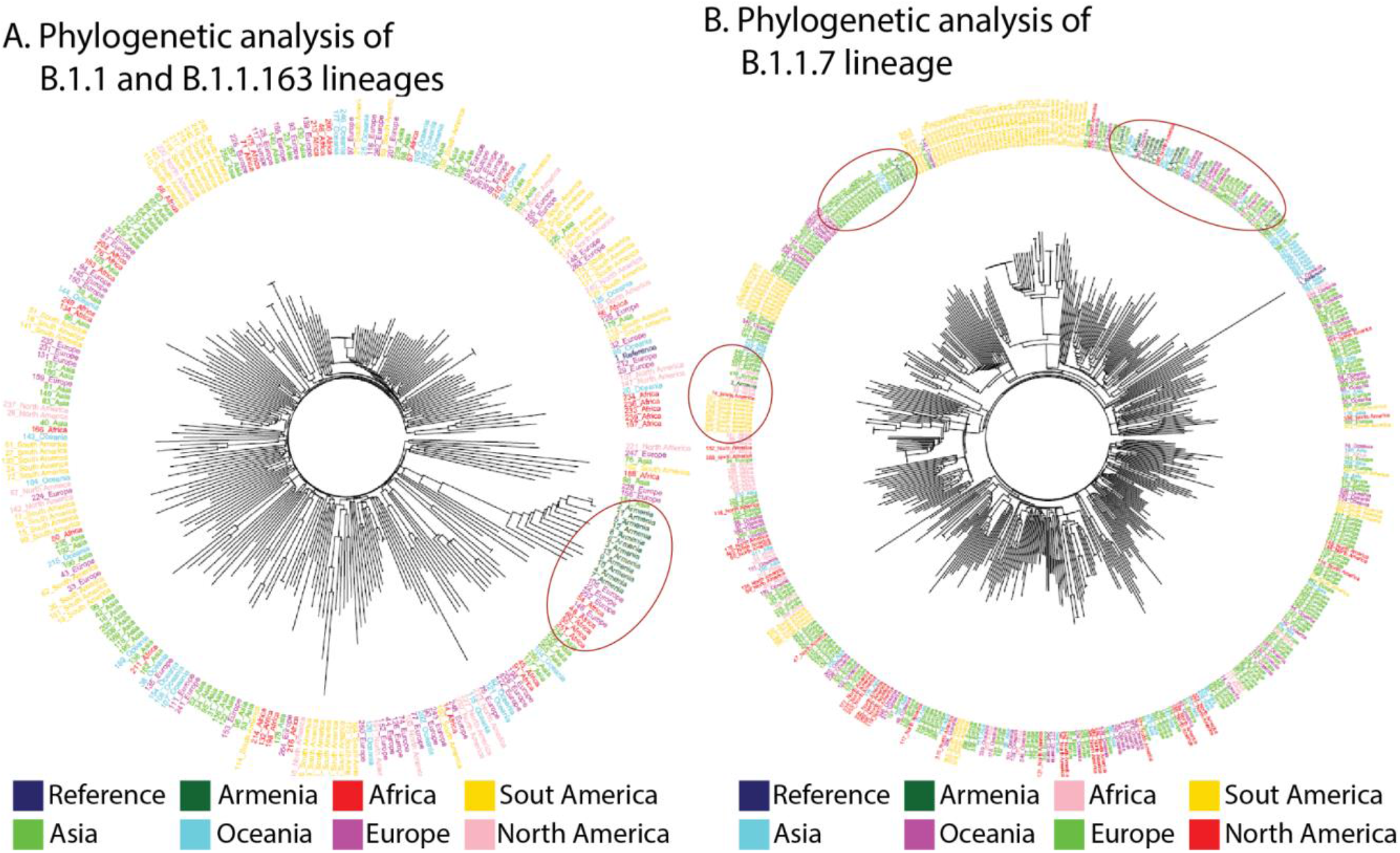
Radial representation of the SARS-CoV-2 phylogenetic tree. Localization of Armenian samples on the phylogenetic trees is indicated by red ovals.

Next, we evaluated the possible effect of identified mutations on the efficacy of recommended primers and probes used for PCR detection of SARS-CoV-2 with COV-GLUE (Table 1).

**Table 1.**
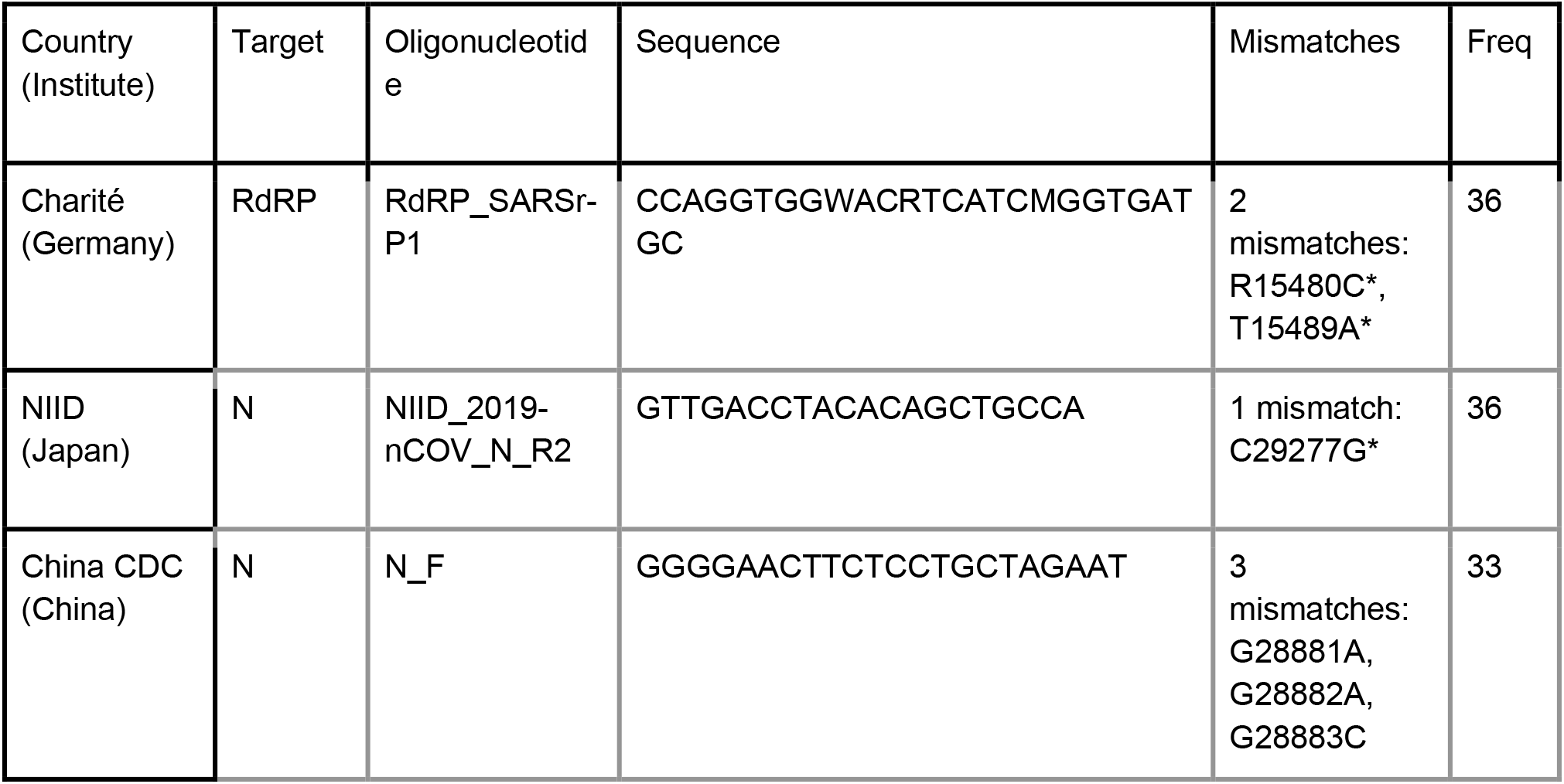
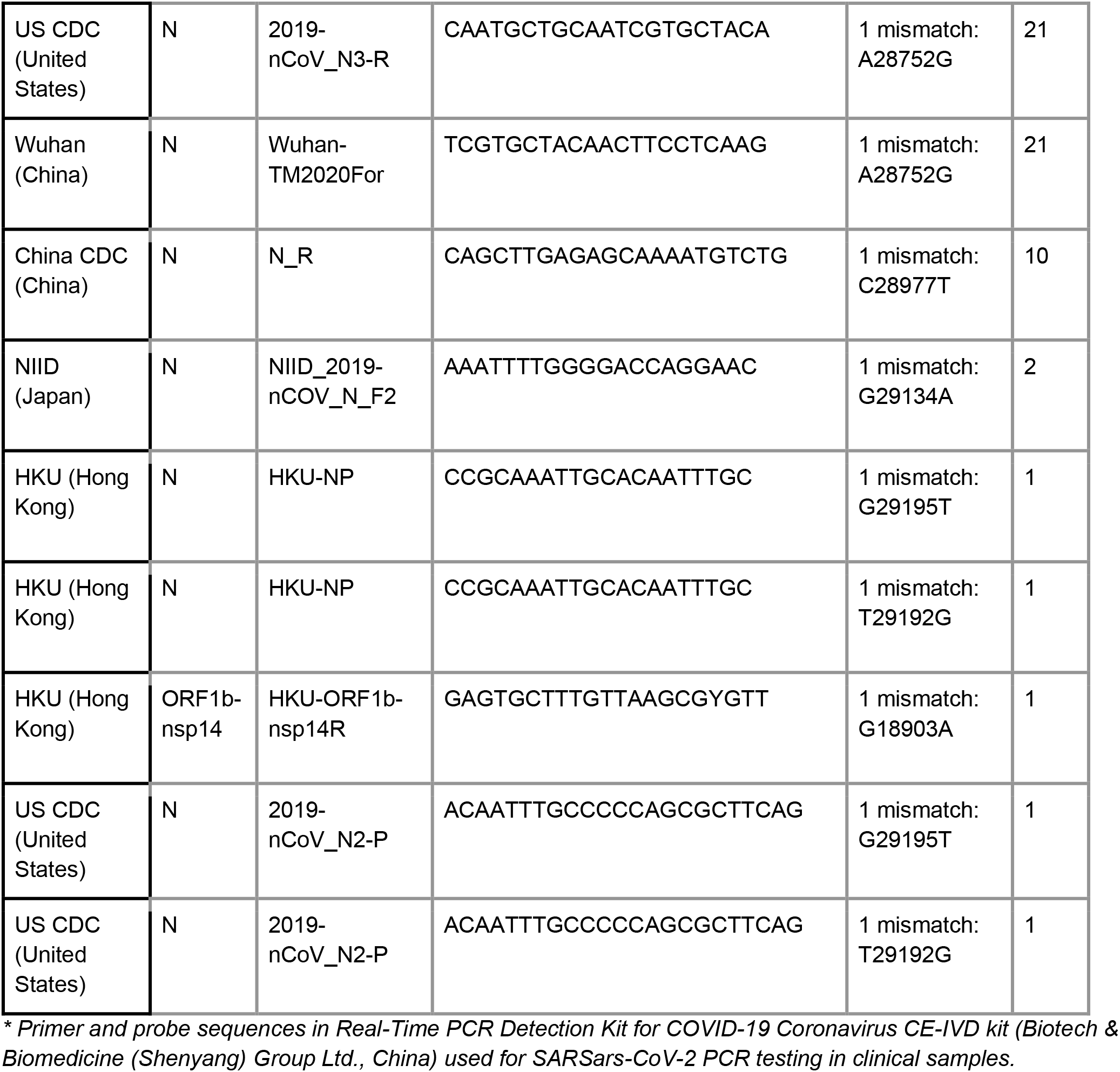
Point mutations in primers and probes for the detection of SARS-CoV-2 of global research institutions *.

The results of the analysis showed that mutations in samples spanned across many regions targeted by primers and probes from different protocols. Regions targeted by Charité primers/probes for RdRP and NIID Japan N gene and US CDC N3 contained the highest number of mutations. N3 forward primer mutations located in 5′ regions and did not influence the primer binding since we obtained the N gene PCR signal in all studied samples (Supplemental Table S1).

## Discussion

COVID-19 outbreak, which started at the end of 2019, is a catastrophic burden to healthcare systems worldwide. As of 2 June 2021; more than 1,800,000 SARS-CoV-2 genome sequences have been deposited in the GISAID EpiCoV database, an open-access global science initiative [27,28]. Nextstrain (https://nextstrain.org) lately designated the new Nextstrain clade 21A and currently divides SARS-CoV-2 diversity into 13 major global clades (19A, 19B, 20A–20J, and 21A), based on high prevalence, signature mutations, and geographic spread. Recently, several lineages (Alpha (UK) B.1.1.7, Beta (South Africa) B.1.351, Gamma (Brazil) P.1, Zeta (Brazil) P.2 and Delta (India) B.1.617.2) have emerged that harbored mutations that increased transmissibility as well as may potentially impair the natural and vaccination-induced immune protection [30]. In this sense, the prompt identification and traceability of disease pathogens and genetic variations are of special importance and interest for disease monitoring and prevention.

In this study, we performed nanopore sequencing of 36 positive samples to perform whole-genome analysis and to characterize SARS-CoV-2. The results showed considerable change in lineage composition throughout September 2020 - March 2021. B.1.1.163 became dominant in January samples. These samples contained characteristic D614G mutation, which is known to increase virion spike density and infectivity [31,32]. Another study also confirmed that D614G substitution activates viral replication in human lung epithelial cells and primary human airway tissues by increasing the infectivity and stability of virions [33]. Among other common mutations identified in the studied samples, C241T was shown to appear with the same frequency as A23403G, C14408T, and C3037T. It has been shown that the C241T mutation changes the C-residue in the SL5b hexameric loop motif of SARS-CoV-2 and is one of the earliest most prevalent (34.8%) SARS-CoV-2 mutation [34]. These mutations co-occur and comprise the major clade G.

The SARS-CoV-2 B.1.1.7 variant is estimated to have emerged in Kent, United Kingdom (UK) in September 2020, and was later classified as a variant of concern (VOC), also referred to as VOC-20DEC-01 [35]. In Armenia B.1.1.7 Alfa lineage first has been detected in clinical samples collected in March. This genetic lineage is suspected to be associated with an increased human-to-human viral transmissibility. This variant has the following lineage-defining non-synonymous mutations: 2 deletions and 6 mutations in spike glycoprotein, an early stop codon and 2 mutations in non-structural protein 8, and 2 mutations in nucleoprotein [36]. From these mutations, N501Y, A570D, and P681H variants in spike glycoprotein thought to affect the structural changes to the properties of it, were found in all our samples [37]. Another mutation, found in our samples, was NS8 Q27stop, an early stop codon in non-structural protein 8. No other mutations assigned to B.1.1.7 lineage were identified.

Besides the molecular genetic characterization of SARS-COV-2 strains, the results helped to understand the transmission routes of the virus to Armenia. B.4 lineage is termed as “Iranian” and was detected in Armenia in September 2020 when the only open border was with Iran. The B.1.1.163 lineage is termed “Russian” because of its high abundance in Russia. In January only incoming flights to Armenia available were from Russia. The phylogenetic analysis confirmed the similarity of B.1.1.163 with Russian lineages, which suggests the penetration of this lineage from Russia. In contrast to that, B.1.1.7 lineages were scattered on the phylogenetic tree and branched with lineages from various geographic regions. These results suggest multiple transmission routes which are in agreement with the relative liberalization of incoming travel restrictions in Armenia in February-March.

Our results also show a higher accumulation of mutations in regions covered by several primers/probes from a clinical perspective. This observation is of particular importance, since mutations may lead to alteration of sensitivity of qPCR tests. Diagnostic tests mostly used in Armenia target ORF1ab and N gene, and our results suggest that identified mutations will not influence their accuracy.

The results of the study again emphasize the need for constant sequencing-based surveillance of SARS-CoV-2 strains for public health decision-making and health care. Nanopore sequencing can serve as an efficient and affordable alternative to short-read next-generation sequencing and be used for epidemiologic surveillance and molecular-genetic analyses of SARS-CoV-2. This is particularly important in countries with underdeveloped NGS sequencing facilities, such as Armenia, and can play an important role in shaping local, national, and regional COVID-19 response strategies.

## Data Availability

Consensus sequences reported in the study were deposited in GISAID EpiCov database.

https://www.gisaid.org/

## Acknowledgments

We would like to thank Vicent Pelechano (SciLifeLab, Department of Microbiology, Tumor and Cell Biology. Karolinska Institutet, Solna, Sweden) for their valuable technical and methodological support.

