## Supplemental Table S1 for "Molecular Genetic Analysis of SARS-CoV-2 Lineages in Armenia"

Supplementary data

**Supplemental Table S1.** The viral RNA load expressed in Ct values performed by qRT-PCR targeting the ORF1ab and N genes in the conserved region of the SARS-CoV-2 genome.

| **Sample ID** | **Ct (ORF1ab gene)** | **Ct (N gene)** |
| --- | --- | --- |
| **1** | 26.25 | 25.52 |
| **2** | 23.72 | 23.06 |
| **3** | 25.44 | 25.54 |
| **4** | 24.65 | 24.45 |
| **5** | 22.27 | 22.09 |
| **6** | 25.56 | 25.67 |
| **7** | 26.25 | 25.69 |
| **8** | 24.17 | 23.55 |
| **9** | 20.36 | 20.37 |
| **10** | 22.18 | 22.41 |
| **11** | 21.25 | 21.15 |
| **12** | 18.98 | 17.97 |
| **13** | 16.56 | 17.92 |
| **14** | 18.75 | 18.86 |
| **15** | 27.89 | 27.85 |
| **16** | 21.51 | 22.38 |
| **17** | 30.92 | 30.30 |
| **18** | 30.11 | 30.91 |
| **19** | 23.80 | 24.66 |
| **20** | 24.86 | 25.73 |
| **21** | 25.41 | 25.18 |
| **22** | 25.45 | 26.38 |
| **23** | 25.33 | 28.20 |
| **24** | 27.02 | 27.89 |
| **25** | 35.03 | 34.10 |
| **26** | 22.33 | 22.51 |
| **27** | 23.84 | 23.94 |
| **28** | 23.72 | 24.41 |
| **29** | 21.17 | 21.54 |
| **30** | 24.92 | 25.94 |
| **31** | 21.03 | 22.19 |
| **32** | 25.16 | 24.80 |
| **33** | 18.53 | 19.11 |
| **34** | 21.54 | 22.52 |
| **35** | 22.27 | 22.99 |
| **36** | 24.58 | 25.19 |
